# Local cortical network stimulation as a concept for focal epilepsy treatment

**DOI:** 10.1101/2023.10.30.23297463

**Authors:** D. van Blooijs, M.D. van der Stoel, G.J.M. Huiskamp, M. Demuru, N.F. Ramsey, F.S.S. Leijten

## Abstract

**Background:** Electrical stimulation therapy for epilepsy patients is applied either to the epileptogenic region or to a larger network (e.g. with deep brain stimulation).

**Objective/hypothesis:** Responses to single pulse electrical stimuli (SPES) reveal potential stimulation sites that target the epileptogenic region for cortical network stimulation therapy.

**Methods:** We applied SPES to ten epilepsy patients who underwent intracranial electrocorticography recordings for pre-surgical evaluation. We detected cortico-cortical evoked potentials (CCEPs) in response electrodes after stimulating other pairs of electrodes, revealing effective connections. We calculated event-related spectral perturbation (ERSP) plots in all response electrodes after stimulating other electrode pairs. We detected interictal epileptic discharges (IEDs) before and after each single pulse and calculated the logarithmic IED ratio.

We analyzed whether power suppression in the ERSP occurred in a response electrode when connected with the stimulus pair. We analyzed whether a larger change in IED ratio was accompanied by power suppression in the response electrode or when this electrode was connected with the stimulus pair.

**Results:** We found that SPES has a neuromodulatory effect measured as: 1) the relationship of a CCEP and power suppression, 2) a larger change in IED rate when a CCEP was present, 3) a decrease in IED rate when power suppression was observed.

**Conclusion(s):** Results suggest that stimulation in an area connected to the epileptogenic region can modulate IEDs in this region. SPES might provide a template for localizing a stimulation site outside the epileptogenic region for electrical stimulation treatment of epilepsy.

**Highlights:**

- Stimulation of an electrode pair can suppress power in an electrode on connected tissue.
- Stimulation of an electrode pair changes IED rate in an electrode on connected tissue.
- A decrease in IED rate was accompanied by power suppression.
- SPES indicates potential stimulation sites for neurostimulation therapy in epilepsy.

## Introduction

Electrical brain stimulation is a relatively new therapy for patients with epilepsy. Stimulation targets, evaluated for epilepsy, fall into two broad categories representing different therapeutic strategies: 1) focal stimulation, intended to directly affect the seizure onset zone ^1^; and 2) global stimulation (e.g. vagal nerve stimulation or deep brain stimulation), where nodes within a larger thalamo-cortical-basal ganglia network are targeted, with the goal of influencing seizure initiation and/or propagation within these pathways ^2^. Focal stimulation is thought to be more effective in suppressing seizure activity than global stimulation ^3^.

Several studies applied focal stimulation with effects ranging from a responder rate of 54% (after two years ^4^) to 73% (after nine years of therapy ^5^) to an overall seizure frequency reduction of 67% ^6^ to more than 80% ^7–10^. These studies applied electrical stimuli in the epileptogenic region, but the optimal stimulation location for neurostimulation therapy is currently undefined ^11^. Perhaps, a local cortical network approach would be beneficial.

The therapeutic effect of neurostimulation may be mediated by specific structural ^11^ or functional networks ^12,13^. When these networks in patients with Parkinson’s disease were compared before and after deep brain stimulation, the networks showed topological reorganization towards the networks measured in healthy controls ^14^. When neurostimulation in epilepsy patients is applied for a longer period of time, functional networks undergo reorganization in patients that respond well to electrical stimulation ^15^. Furthermore, the fact that seizure frequency decreases over time when neurostimulation therapy is effective ^5,6^ also supports the idea that networks undergo plasticity-related changes resulting in a network that is less prone to evolving seizures ^15^. If the potential capability of plasticity-related changes could be measured in the individual patient prior to neurostimulation treatment, this would help in decision making towards a personalized therapy.

Analyses of electrocorticography (ECoG) recordings have shown that the degree of synchronizability of the network could predict the effectiveness of neurostimulation treatment ^16^. Another study shows that stimulation in the epileptogenic region was more effective in seizure rate reduction if the node had more connections with other nodes ^17^. This suggests that it is important to also look at the underlying network to determine the optimal stimulation site.

An effective network can be derived from single pulse electrical stimulation (SPES) ^18^ in which single pulses are applied to intracranial electrode pairs. Cortico-cortical evoked potentials (CCEPs) to these stimuli in other electrodes indicate a connection between the stimulus pair and the responding electrode ^18^. The epileptogenic tissue was found to exhibit a higher density of such connections than surrounding tissue ^19^. We hypothesize that electrical stimulation in a stimulus pair connected to the epileptogenic cortical network might facilitate plasticity-related changes as described in long-term electrical stimulation studies.

In this study, we investigate whether the existence of an effective connection (by means of an evoked CCEP) between the stimulus pair and a responding electrode facilitates stimulation-induced changes in rate of interictal epileptic discharges (IEDs), and whether it can affect neural activity in terms of spectral power changes. Effects on IEDs and neural activity in general could be of interest as a surrogate marker ^20^ for electrical stimulation therapy in that it could offer new stimulation target options.

## Materials and methods

### Patient characteristics

We selected epilepsy patients who underwent electrocorticography (ECoG) recordings for 5-7 days for evaluation of epilepsy surgery between 2014 and 2017. We decoded and visually annotated the data for bad channels, artefacts, seizures and stimulus pairs, and imported this data into the Brain Imaging Data Structure (BIDS) ^21,22^. The study was performed with approval from the ethical committee under Dutch law, in accordance with the Declaration of Helsinki (2013).

### Acquisition and pre-processing epochs

A Single Pulse Electrical Stimulation (SPES) protocol had been performed in these patients as part of clinical routine to delineate epileptogenic tissue ^23^. Ten monophasic pulses (8 mA, 1 ms, 0.2 Hz) were applied to pairs of adjacent electrodes (Figure 1). In the primary sensorimotor cortex, pulse intensity was decreased to 4 mA to avoid tingling or twitches. Electrodes located on top of other electrodes or electrodes with noisy signals were not stimulated and excluded from analysis.

**Figure 1:**
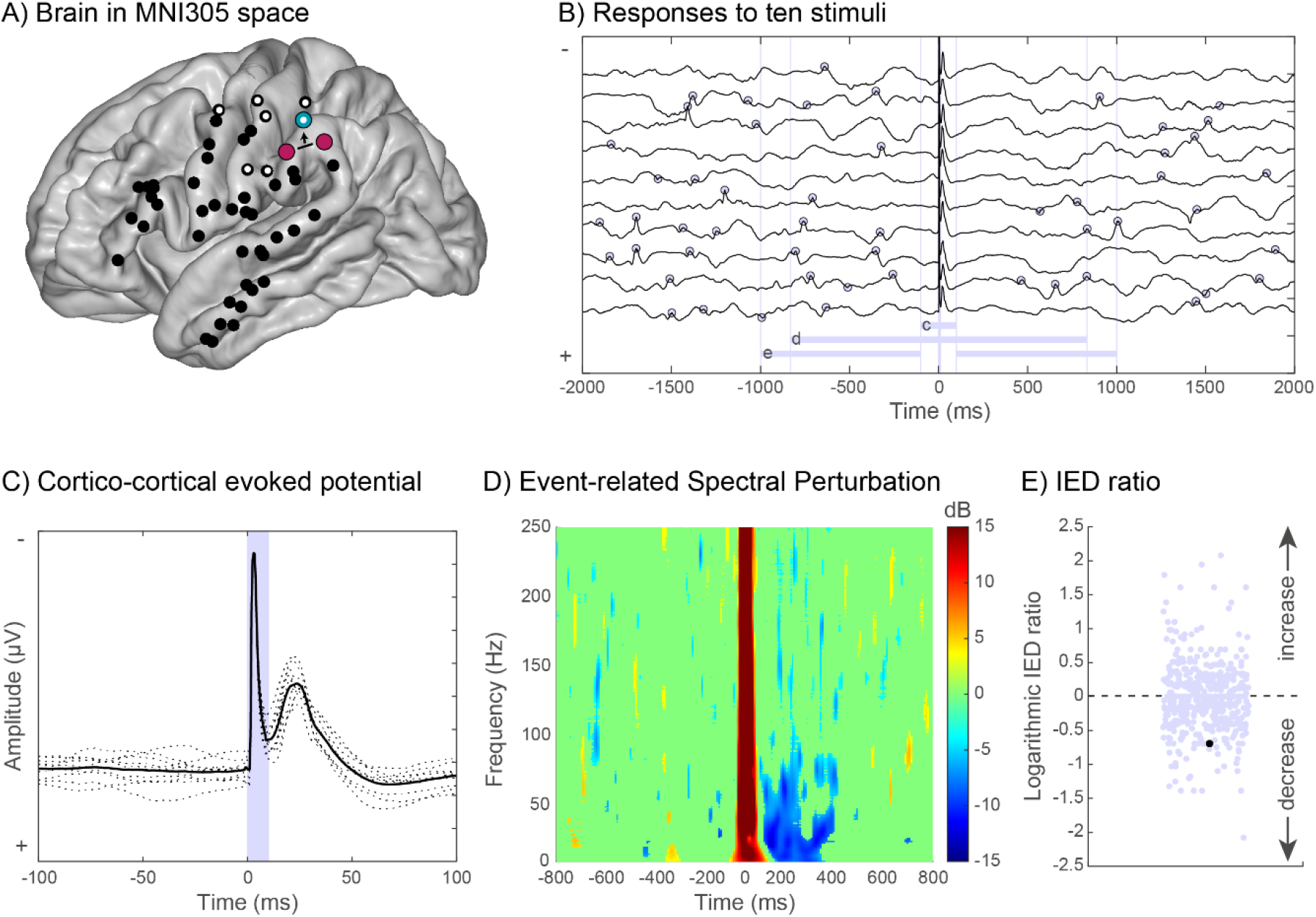
overview of one patient. A) the grid electrodes in MNI305 space. The electrodes with a white dot are the electrodes in which IEDs were observed. The purple electrodes are stimulated and the responses in the blue electrode are shown in B). In B), ten responses of 2 s pre- and 2 s post-stimulation are displayed. The gray dots indicate the detected IEDs. We observed that the number of IEDs seemed to be reduced during 500 ms after stimulation. The time windows in which respectively CCEPs, power suppression in ERSP, and IED ratio are determined are visualized with gray bars at the bottom of this figure. C) Ten single cortico-cortical evoked potentials (CCEPs) (dotted lines) of B) and the average response (black line) are visualized in a time window of 100 ms pre- and 100 ms post-stimulation. The peak at ∼30ms after stimulation is called the N1 peak and is the first negative deflection that is characteristic for the CCEP. The gray vertical bar between 0 and 10 ms displays the time window in which the stimulus artefact is visible. D) An Event-Related Spectral Perturbation (ERSP) plot is constructed based on the ten epochs in B). The red bar at 0 ms indicates the stimulation artefact. The blue area after the stimulation artefact indicates a significant suppression in power compared to the time window pre-stimulation. This suppression is present in the frequency band from 1 to ∼100 Hz during 400 ms after stimulation. E) The logarithmic IED ratios of this specific subject are displayed in gray. The black dot indicates the logarithmic IED ratio derived from the ten epochs in B). During 1 s pre-stimulation, 14 IEDs were detected, during 1 s post-stimulation 7 IEDs were detected, resulting in a logarithmic IED ratio of -0.69.

For each electrode, ten epochs in time domain were averaged for each stimulation pair, with a time window of 2 s before to 2 s after the electrical stimulus time-locked to the stimulus (Figure 1B). We detected cortico-cortical evoked potentials (CCEPs, Figure 1C) when the first negative deflection within 100 ms after stimulation exceeded a threshold of 2.6 times the standard deviation that was calculated in an epoch interval of -2s to -0.1s covering pre-stimulus baseline ^19,24^. The detected CCEPs were visually checked (DvB).

For each electrode, a wavelet-based time-frequency transformation was applied to ten epochs with a time window of 1 s before to 1 s after the stimulus time-locked to the stimulus for each stimulation pair ^25^. This Event-Related Spectral Perturbation (ERSP ^26^) plot used a Morlet wavelet with two oscillation parameters [3 0.8]. The frequency range was set to 10-250 Hz with a frequency resolution of 1 Hz. We applied bootstrapping to observe significant spectral changes post-stimulation compared to pre-stimulation (Figure 1D). We used a trained support vector machine, based on the surface, the duration and the frequency range of an area with power suppression, to detect these significant events of power suppression post-stimulation. The ERSPs with detected power suppression were visually checked (DvB).

### Preprocessing interictal epileptic discharges (IEDs)

For each patient, electrodes showing interictal epileptic discharges (IEDs) were determined by a clinical neurophysiologist (FL). We applied an IED detection algorithm based on the algorithm by Gaspard et al. 2014 ^27^ to the time domain recordings in which the SPES protocol was executed. In each electrode with IEDs, we counted the number of IEDs 1-0.1 s before and 0.1-1 s after stimulation for each individual stimulus. We excluded a symmetric time window of in total 200 ms around the stimulus onset to exclude CCEPs which occur within 100 ms after stimulation and to avoid interference of the stimulus artefact in counting the number of IEDs (Figure 1B).

We calculated the ratio (Figure 1E) by dividing the number of IEDs post-stimulation by the number of IEDs pre-stimulation across the ten pulses to each stimulus pair. We converted the ratio to a logarithmic scale: a value of 0 means that no change was observed in number of IEDs post-stimulation compared to pre-stimulation; a value of <0 means a decrease in IEDs after stimulation; and a value of >0 means an increase in IEDs after stimulation.

### Analysis

First, we investigated whether CCEPs in a response electrode were accompanied by power suppression in the ERSP after stimulating a certain electrode pair by calculating the odds ratio for all individual patients and all patients combined. Odds ratios were analyzed with chi^2^ test and FDR corrected (p<0.05). For this analysis, we included all subdural electrodes.

For the following analyses, we only included the electrodes in which IEDs were observed. We investigated whether the occurrence of a CCEP was accompanied by a change in IEDs post-stimulation. We separated IED electrodes with and without a CCEP and analyzed whether the distribution of IED ratios was statistically different with a Kolmogorov-Smirnov test. If both distributions differed, we analyzed this difference in more detail. We compared the absolute values of logarithmic IED ratio, defined as either increase or decrease in IED ratio, to explore general changes in neuromodulation that might be induced when an IED electrode is connected with the stimulus pair. We also compared the positive values of logarithmic IED ratio, which means only an increase in IEDs post-stimulation, and the negative values of logarithmic IED ratio, which means only a decrease in IEDs post-stimulation to investigate whether a specific effect of neuromodulation was induced when an IED electrode was connected to the stimulus pair. Comparisons in logarithmic IED ratios were analyzed with the Mann Whitney U test and FDR corrected (p<0.05). We also separated IED electrodes with and without the occurrence of power suppression after stimulation of stimulus pairs and repeated the statistical analyses on IED ratios as described earlier.

Finally, we investigated the number of IEDs in time. We wanted to analyze how long a change in IED count post-stimulation compared to pre-stimulation would last. Therefore, we counted the number of IEDs in an epoch of 2 s pre- and 2 s post-stimulation instead of 1 s pre- and post-stimulation, as was used for the logarithmic IED ratio. We categorized response electrodes into four categories: 1) with evoked CCEP and power suppression, 2) with evoked CCEP and without power suppression, 3) without CCEP and with power suppression, 4) without CCEP and without power suppression. We calculated how many IEDs were observed on average during the 2 s pre-stimulation. We divided the 2 s post-stimulation time window in periods of 200 ms and calculated how many IEDs were observed on averaged in each period for each category. We compared the number of IEDs in each consecutive time window of 200 ms post-stimulation with the mean number of IEDs pre-stimulation. Comparison in number of IEDs in consecutive time windows were analyzed with a paired t-test and FDR corrected (p<0.05).

### Code and data availability

All code is available at https://github.com/dvanblooijs/CCEP_suppressionPower_Spikes. Data will be available at openneuro.org when the study is published in a peer-reviewed journal.

## Results

### Patient characteristics

We included ten patients (6 males, median age 15 years (range: 9-41 years), see Table 1). ECoG was performed with subdural platinum circular electrodes with 4.2 mm^2^ contact surface, and an center-to-center electrode distance of 1 cm (AdTech). Electrode grids were placed on the brain areas suspected of seizure onset. A median number of 64 (range: 54-96) electrodes were implanted per patient. In four patients, depth electrodes (1-2 leads with 6 electrodes each) were placed in the presumed lesion that was visible on MRI. Data were recorded with a sampling frequency of 2048 Hz. In seven patients, a median number of 8 electrodes (range: 6-19) were covering areas generating IEDs. In three patients, no IEDs were observed. These three patients (5, 7, 9) were excluded when analyzing the IED ratio.

**Table 1:** patient characteristics. M = male, F = female, T = temporal, Oc = Occipital, F = Frontal, C = Central, IH = interhemispheric, D = depth electrode in presumed lesion on MRI, P = Parietal, R = right, L = left.

| Patient # | Sex | location | Side | # stimulated electrode pairs | # electrodes | # electrodes with IEDs |
| --- | --- | --- | --- | --- | --- | --- |
| 1 | M | T, Oc | R | 45 | 64 | 8 |
| 2 | F | F | L | 55 | 64 | 19 |
| 3 | F | F, C, IH | L | 70 | 80 | 6 |
| 4 | M | F, IH, D | L | 53 | 62 | 8 |
| 5 | F | T | L | 44 | 64 | - |
| 6 | M | F, D | L | 43 | 60 | 13 |
| 7 | M | T, P | L | 69 | 96 | - |
| 8 | M | T, C, D | L | 47 | 54 | 10 |
| 9 | F | C, D | R | 58 | 70 | - |
| 10 | M | C | R | 56 | 64 | 8 |

The odds ratio for the occurrence of an evoked CCEP accompanied by power suppression when stimulating a specific stimulus pair was between 4.7 (CI: 3.0-7.4) and 11.4 (CI: 10-13) for all individual patients and 8.0 (CI: 7.5-8.5) when these patients were combined (Figure 2).

**Figure 2:**
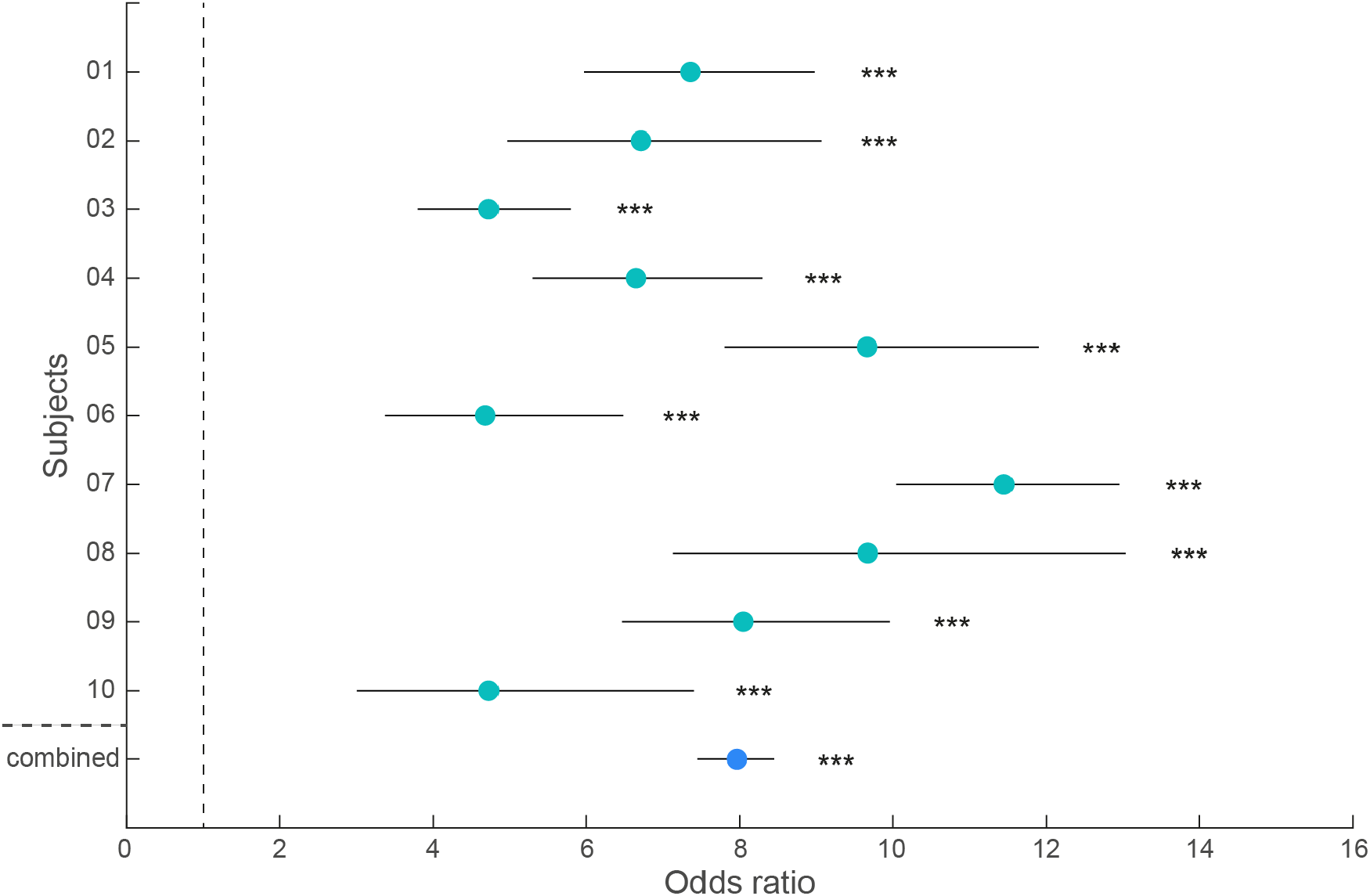
The occurrence of a CCEP accompanied by power suppression in a response electrode when stimulating a specific stimulus pair is displayed for all individual patients and for all patients combined. The odds ratio varied between 4.7 (CI: 3.0-7.4) and 11.4 (CI: 10-13) in individual patients and was 8.0 (CI: 7.5-8.5) in all patients combined. *** = p<0.001, FDR corrected.

When we compared the IED ratio in response electrodes with or without evoked CCEP after stimulating a specific stimulus pair, the distributions differed significantly (Figure 3A, p<0.01). The absolute values and positive values of IED ratio were increased in response electrodes accompanied by an evoked CCEP (Figure 3B-C). We also observed a decrease in negative values of IED ratio in response electrodes accompanied by an evoked CCEP (Figure 3D).

**Figure 3:**
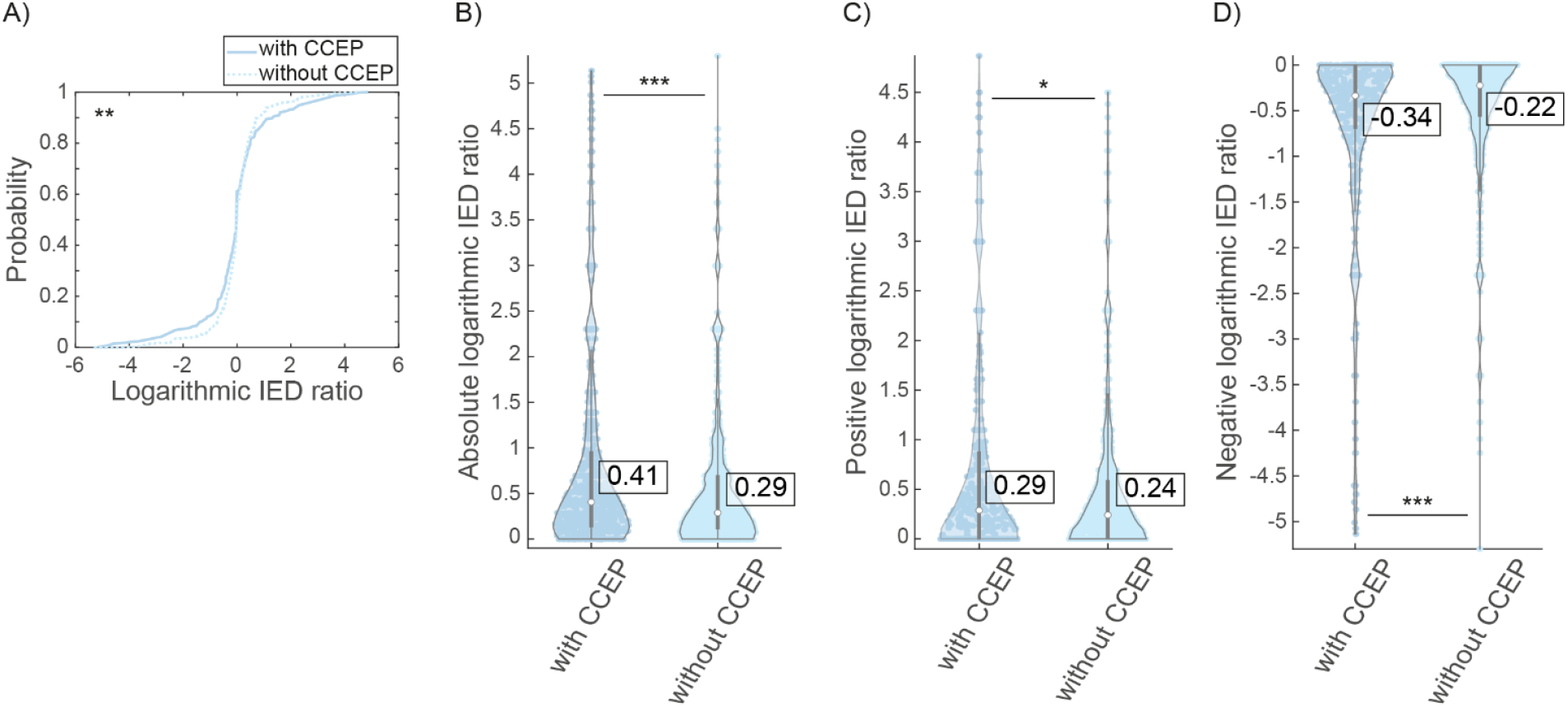
Logarithmic IED ratios in response electrodes when a CCEP was (not) evoked after stimulating other electrode pairs. A) The cumulative IED ratio, showing that there was a significant difference in distributions of Logarithmic IED ratios when a CCEP was (not) evoked. B) The absolute values of logarithmic IED ratio, indicating a larger change in IEDs after stimulation when a CCEP was evoked. C) The positive values of logarithmic IED ratio, indicating a larger increase in number of IEDs after stimulation when a CCEP was evoked. D) The negative values of logarithmic IED ratio, indicating a larger decrease in number of IEDs after stimulation when a CCEP was evoked. * = p < 0.05, ** = p<0.01, *** = p<0.001, FDR corrected.

When we compared the IED ratio in response electrodes with or without power suppression in the ERSP after stimulating a specific stimulus pair, the distributions differed significantly (Figure 4A, p<0.001). The absolute values of logarithmic IED ratio was increased when power suppression was observed in a response electrode (Figure 4B). We did not find any difference in positive values of IED ratio (Figure 4C), but we observed a decrease in negative values of IED ratio when power suppression was observed in a response electrode (Figure 4D).

**Figure 4:**
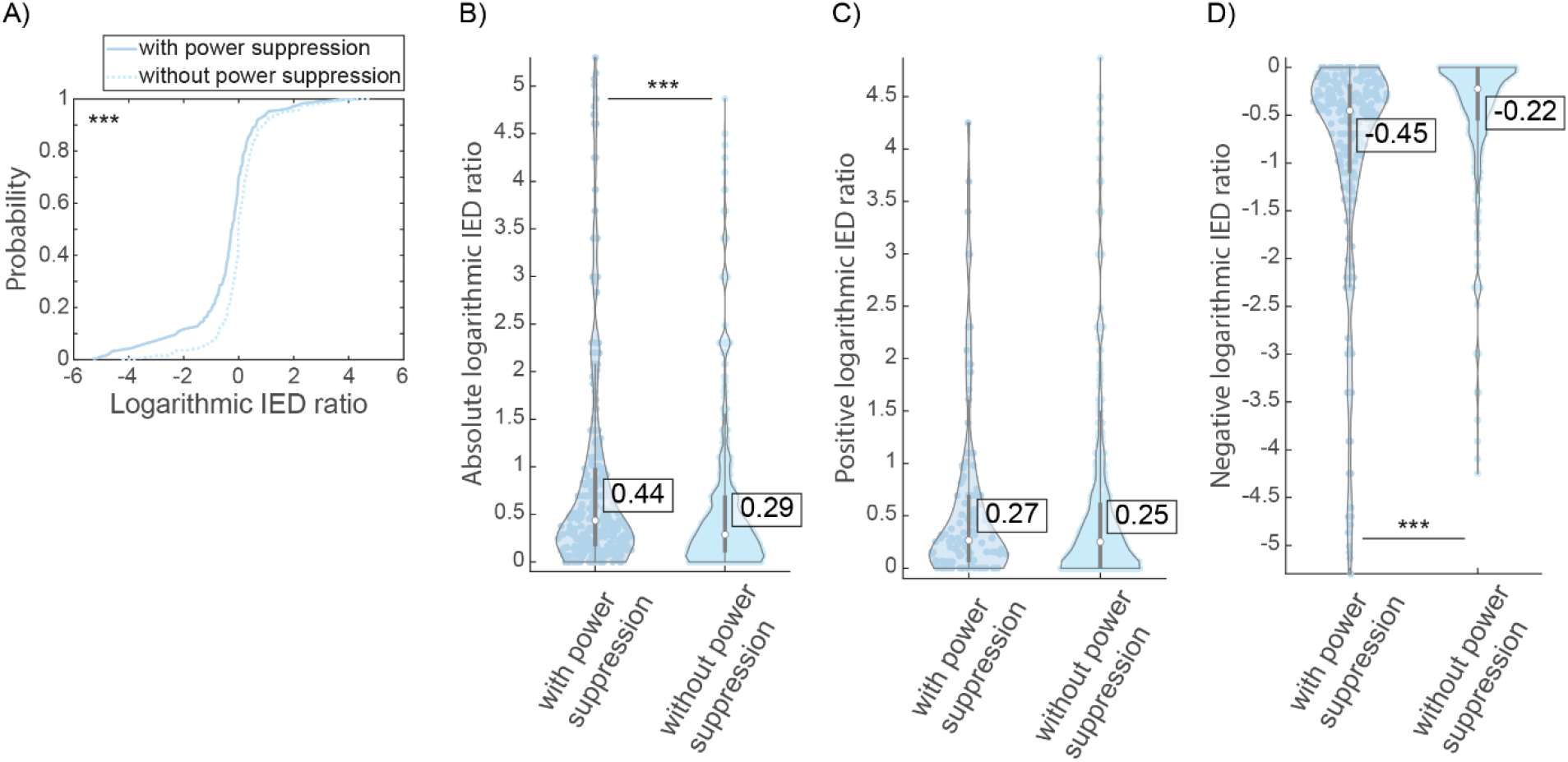
Logarithmic IED ratios in response electrodes when power suppression was (not) observed after stimulating other electrode pairs. A) The cumulative IED ratio, showing that there was a significant difference in distributions of logarithmic IED ratios when power suppression was (not) observed. B) The absolute values of logarithmic IED ratio, indicating a larger change in IEDs after stimulation when power suppression was observed. C) The positive values of logarithmic IED ratio, no difference was found when power suppression was (not) observed. D) The negative values of logarithmic IED ratio, indicating a larger decrease in number of IEDs after stimulation when power suppression was observed. *** = p<0.001, FDR corrected.

From seven subjects combined, we categorized IED electrodes after stimulating stimulus pairs into four categories: 1) with evoked CCEP and power suppression (n=169), 2) with evoked CCEP and without power suppression (n = 517), 3) without CCEP and with power suppression (n = 175), 4) without CCEP nor power suppression (n = 2741).

When we analyzed if and how long IEDs were affected by SPES stimuli (Figure 5), we observed that the numbers of IEDs were decreased post-stimulation when accompanied by power suppression and/or a CCEP (Figure 5A, C, D, G). This decrease in number of IEDs was most pronounced in electrodes that showed power suppression, regardless of the presence of a CCEP (Figure 5G, 0.2-0.4, 0.6-1, 1.2-1.4, 1.6-1.8 s post-stimulation).

**Figure 5:**
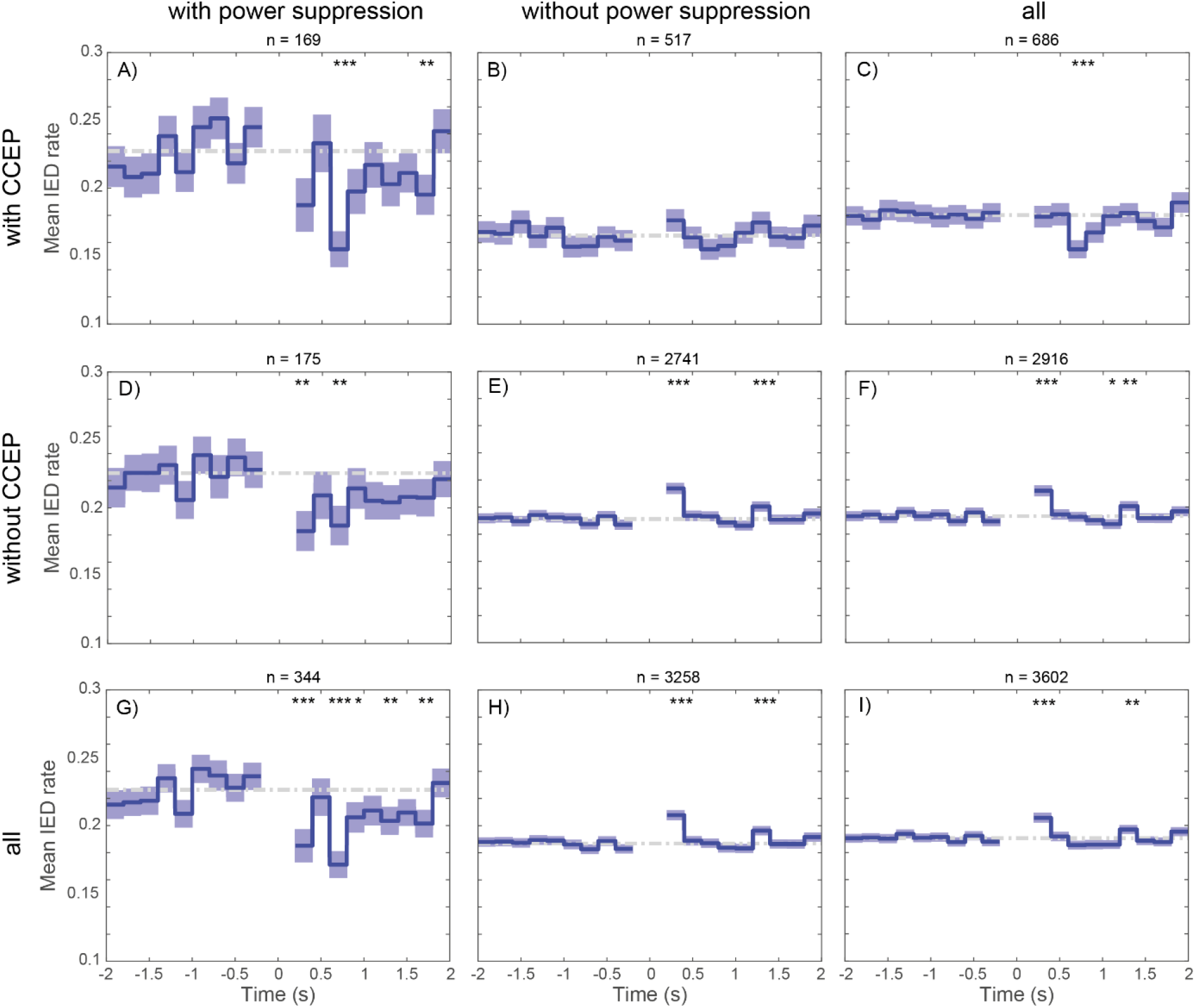
block signals displaying the mean number of IEDs and standard error of the mean in consecutive periods of 200 ms. A time window of 400 ms around the stimulus artefact (t = 0 s) was excluded from analysis. Numbers of IEDs in each consecutive period of 200 ms post-stimulation were compared with the mean number of IEDs pre-stimulation (gray dotted line). A) number of IEDs when both a CCEP and power suppression were observed in the response electrode. There was a decrease in number of IEDs between 0.6-0.8 s and 1.6-1.8 s post-stimulation. B) number of IEDs when a CCEP and no power suppression was observed. There was no change in number of IEDs post-stimulation. C) number of IEDs when a CCEP was observed, regardless of the presence of power suppression. There was a decrease in number of IEDs between 0.6-0.8 s after stimulation. D) number of IEDS when power suppression and no CCEP was observed. There was a decrease in IEDs 0.2-0.4 s and 0.6-0.8 s after stimulation. E) number of IEDs when no power suppression or CCEP was observed. There was an increase in IEDs 0.2-0.4 s and 1.2-1.4 s after stimulation. F) number of IEDs when no CCEP was observed, regardless of the presence of power suppression. There was an increase in IEDs 0.2-0.4 s and 1.2-1.4 s after stimulation, but a decrease in IEDs 1.0-1.2 s after stimulation. G) number of IEDs when power suppression was observed, regardless of the presence of a CCEP. There was a decrease in IEDs 0.2-0.4 s, 0.6-1.0 s, 1.2-1.4 s and 1.6-1.8 s after stimulation. H) number of IEDs when no power suppression was observed, regardless of the presence of a CCEP. There was an increase in IEDs 0.2-0.4 s and 1.2-1.4 s after stimulation. I) number of IEDs in all electrodes, regardless of the presence of a CCEP or power suppression. There was an increase in IEDs 0.2-0.4 s and 1.2-1.4 s after stimulation. *** = p<0.001, ** = p<0.01, * = p<0.05, FDR corrected.

We also observed an increase in number of IEDs 0.2-0.4 s and 1.2-1.4 s after stimulation when there was no power suppression and/or CCEP observed in the response electrode (Figure 5E, F, H), and when all electrodes were combined, regardless of occurrence of CCEP or power suppression (Figure 5I). In IED electrodes with a CCEP but not accompanied by power suppression, we did not see any change in number of IEDs post-stimulation (Figure 5B).

## Discussion

This study provides proof of principle in demonstrating that changes in brain signals are induced by SPES. We found a high association between an evoked CCEP and power suppression in ten individual patients and when these patients were combined. One study^28^ showed that the stimulation response was stronger and exhibited progressive modulation in areas highly connected to the stimulation site. A few other studies ^10,29,30^ mentioned that cortical stimulation outside the epileptogenic region did not have any effect on IED rate. However, they did not investigate underlying effective networks ^10,30^, or no effective connection between the stimulus pair outside the epileptogenic region and the electrodes in the epileptogenic region was observed ^29^.

We found a larger absolute, positive and negative value in logarithmic IED ratios when a response electrode was connected to the stimulus pair, indicating that both an increase as well as a decrease, which was more pronounced, in number of IEDs could occur. Traditionally, IEDs are assumed to represent short bursts of seizure activity, but without becoming clinical seizures ^31^. Another hypothesis is that IEDs increase the threshold for a seizure to occur which means that IEDs would have a protective function ^32^. Whether IEDs have a facilitating or preventive function for seizures might even depend on the dynamical state of the brain ^33^. The clinical implications of the two interpretations of IEDs are quite contradictory, leading to discussions whether you would like to suppress this activity with electrical stimulation. Alarcon et al. ^31^ found similarities in neuronal firing patterns associated with IEDs and SPES and conclude that a period of suppression in firing pattern does not result from the intrinsic properties of membranes but from the properties of the neuronal network. In the current study, we assume that both an increase and a decrease in IED rate is an indication that stimulation at a specific site has a modulating effect on epileptic activity. Further research with varying stimulus parameters in long-term electrical stimulation should give more insight in whether increase or decrease of IED rate is a good surrogate marker for effective stimulation therapy.

We also observed a decrease in IED ratio when the response electrode showed power suppression, which means that the number of IEDs after stimulation was reduced. The phenomenon of power suppression after SPES has been described in two studies ^34,35^. These studies only looked at power suppression in high frequencies (>70 Hz). They both conclude that power suppression was significantly stronger in the epileptogenic tissue, but Maliia et al.^35^ also found power suppression in the default mode network during 0.2-0.5 s after stimulation. Our data showed that power suppression in the response electrode would be observed more often when a direct connection, indicated by a CCEP, between the stimulus pair and the response electrode was present. This observation was not limited to the electrodes covering epileptogenic tissue and was present when including all implanted electrodes in this analysis, suggesting that it is not a direct mathematical effect of decreased number of IEDs.

When power suppression was observed in a response electrode, a decrease in IED rate after stimulation was visible between 0.6-1.8 s after stimulation, although this was not significant during this whole period. The power suppression in ERSP plots typically occurred within a time window of 0.2-0.4 s after stimulation (Figure 1D), which means that the actual decrease in IED rate had a longer duration than was visualized in these ERSP plots. Stypulkowski et al. ^37^ investigated whether stimulus-induced reduction in activity was associated with reduced excitability and found that after-discharges were almost completely blocked and the amplitude of evoked potentials was reduced. Keller et al. ^38^ used the ratio of high-amplitude CCEPs, before versus after applying repetitive stimulation trains of 10 Hz, as a measure of cortical excitability and found that the CCEP-amplitude was modulated following repetitive stimulation. This suggests that the changes in brain signals after SPES itself, namely power suppression and the change in IED rate after stimulation, could be an interesting measure of cortical excitability. Since cortical excitability is increased for several hours before a seizure occurs ^41^, and many anti-epileptic drugs affect neural excitability to reduce the risk of seizures ^42^, the power suppression after SPES could be of importance in localizing optimal stimulation sites for effective stimulation therapy.

A striking observation was that when no power suppression or CCEP was present in an electrode, an increase in IED rate was observed between 0.2-0.4 s and 1.2-1.4 s after stimulation. Delayed responses, spikes or sharp waves occurring between 0.1-1 s after SPES ^39^, are represented as power increase in ERSP plots, and are a biomarker for epileptogenic tissue ^25^. This increase in IEDs is found in electrodes that were not connected to the stimulus pair by a CCEP, which supports the observation that these delayed responses occur more often in indirect connections ^40^.

This preliminary investigation is limited to a small group of patients with heterogeneous ECoG coverage based on clinical evaluation of suspected epileptogenic tissue. It was not possible to record responses from the stimulated electrodes because of large stimulation artifacts or saturation of the amplifier that lasted for 4-5 s. Therefore, we did not have the possibility to compare responses to local stimulation in epileptogenic tissue with the observed responses to cortical network stimulation as described in this study.

Several studies ^9,10^ investigated whether electrical stimulation affected the IED frequency before they implanted a neurostimulator. Unfortunately, time is limited during an intracranial monitoring period and it is not possible to test the great variety of stimulation parameter combinations in several stimulation sites. Especially in large epileptogenic regions, the optimal effect of stimulation might not be observed due to a suboptimal stimulation site, or suboptimal stimulus parameters, missing the potential that electrical stimulation might have for the specific patient. In this study, we used SPES to probe the brain in all locations covered by ECoG which gave us an indication of potential stimulation sites that might be beneficial for patient-tailored cortical stimulation therapy to reduce seizure frequency.

In conclusion, we found stimulus-induced neuromodulatory effects, by means of change in IED rate and change in spectral power, when SPES was applied in a response electrode connected to the stimulus pair. This could have a great potential to select stimulation sites for cortical network stimulation therapy.

## Acknowledgements

This project was funded by the Dutch Epilepsy Fund (NEF 17-07). We thank S. Blok for reading the manuscript. We thank the SEIN-UMCU RESPect database group (C.J.J. van Asch, L. van de Berg, S. Blok, M.D. Bourez, K.P.J. Braun, J.W. Dankbaar, P. van Eijsden, C.H. Ferrier, T.A. Gebbink, R. van Griethuysen, M.G.G. Hobbelink, F.W.A. Hoefnagels, N.E.C. van Klink, M.A. van ‘t Klooster, G.A.P. de Kort, M.H.M. Mantione, A. Muhlebner, J.M. Ophorst, P.C. van Rijen, S.M.A. van der Salm, E.V. Schaft, M.M.J. van Schooneveld, H. Smeding, D. Sun, A. Velders, M.J.E. van Zandvoort, G.J.M. Zijlmans, E. Zuidhoek, W.J.E.M. Zweiphenning, J. Zwemmer) for their contributions and help in collecting the data.

